# Staffing and Capacity Planning for SARS-CoV-2 Monoclonal Antibody Infusion Facilities: A Performance Estimation Calculator based on Discrete-Event Simulations

**DOI:** 10.1101/2021.07.23.21260984

**Authors:** Çağlar Çağlayan, Jonathan Thornhill, Miles A. Stewart, Anastasia S. Lambrou, Donald Richardson, Kaitlin Rainwater-Lovett, Jeffrey D. Freeman, Tiffany Pfundt, John T. Redd

## Abstract

**Objective:** The COVID-19 pandemic has significantly stressed healthcare systems. The addition of monoclonal antibody (mAb) infusions, which prevent severe disease and reduce hospitalizations, to the repertoire of COVID-19 countermeasures offers the opportunity to reduce system stress but requires strategic planning and use of novel approaches. Our objective was to develop a web-based decision-support tool to help existing and future mAb infusion facilities make better and more informed staffing and capacity decisions.

**Materials and Methods:** Using real-world observations from three medical centers operating with federal field team support, we developed a discrete-event simulation model and performed simulation experiments to assess performance of mAb infusion sites under different conditions.

**Results:** 162,000 scenarios were evaluated by simulations. Our analyses revealed that it was more effective to add check-in staff than to add additional nurses for middle-to-large size sites with ≥ 2 infusion nurses; that scheduled appointments performed better than walk-ins when patient load was not high; and that reducing infusion time was particularly impactful when load on resources was only slightly above manageable levels.

**Discussion:** Physical capacity, check-in staff, and infusion time were as important as nurses for mAb sites. Health systems can effectively operate an infusion center under different conditions to provide mAb therapeutics even with relatively low investments in physical resources and staff.

**Conclusion:** Simulations of mAb infusion sites were used to create a capacity planning tool to optimize resource utility and allocation in constrained pandemic conditions, and more efficiently treat COVID-19 patients at existing and future mAb infusion sites.

## 1. Background and Significance

Spreading rapidly, Coronavirus disease 2019 (COVID-19) became a global pandemic in early 2020 [1]. Caused by severe acute respiratory syndrome coronavirus 2 (SARS-CoV-2), COVID-19 is a highly contagious disease that can result in mild to severe symptoms, hospitalization, need for intensive care or ventilator treatment, and mortality [2]. By July 2021, SARS-CoV-2 had infected more than 33 million Americans, caused over 600,000 deaths in the United States (U.S.) [3], and killed more than 3,900,000 people worldwide [4].

In November 2020, the Food and Drug Administration issued emergency use authorizations for monoclonal antibody (mAb) monotherapy bamlanivimab and combination therapy casirivimab/imdevimab to treat COVID-19 among individuals at high risk for progressing to severe disease [5]. These mAb treatments were shown to be effective for preventing progression of disease and COVID-19-associated hospitalizations [6, 7]. Given that a significant number of individuals remain at risk for COVID-19 in the U.S. [8], mAb treatments will continue to be critical for saving lives and reducing COVID-19 morbidity and mortality [9, 10].

In the context of capacity and staffing shortages, mAb therapies are especially important to reduce burdens on the U.S. healthcare system. However, these treatments have administration challenges, including authorized use only in confirmed COVID-19 outpatients; the need to treat patients as early as possible; and the messaging required to introduce mAb products to providers and patients in an already-stressed system [11]. Moreover, an outpatient mAb treatment requires the allocation of staff, physical space, equipment, and supplies, and involves several sequential treatment procedures necessitating synchronization. Thorough analysis and careful planning are needed to decrease barriers to implementation and ensure efficient service.

The objective of this study was to create a capacity and staffing planning tool to support the implementation of mAb drug products at U.S. healthcare facilities. Taking a simulation approach, we developed a web-based calculator to investigate the operational performance of mAb infusion sites as a function of staffing, capacity, and other key factors. The decision-support tool can be used to identify bottlenecks in the mAb infusion process, and help decision-makers make informed resource allocation decisions for mAb treatment service.

## 2. Materials and Methods

Discrete-event simulation (DES) experiments were conducted for 162,000 alternative scenarios considering different staffing and capacity levels, scheduling protocols, patient demand, facility service hours, and infusion durations. To inform the model structure and collect data for simulation experiments, the mAb treatment process was observed at three U.S. medical centers implementing mAb infusions in collaboration with Disaster Medical Assistance Teams deployed by the US Department of Health and Human Services under the direction of the Assistant Secretary for Preparedness and Response’s National Disaster Medical System. To make simulation results accessible, a web-based decision-support tool was developed, which displays estimated performance outputs for the scenarios associated with user-selected inputs. The DES model and simulation experiments were programmed using Matlab version R2020a.

### Patient Arrival Process

Infusion facilities either had walk-in encounters, where patients were admitted to check-in area on a first-come-first-served basis, or utilized scheduled appointments of three different types: (i) block, (ii) spread-out, and (iii) mixed (**Table 1**). A block schedule uses only a few scheduling points (e.g., 9 AM, 1 PM, 4 PM) and books a batch of patients for each time block. Spread-out scheduling uses numerous scheduling points and books a small number of patients for the same time point to more evenly distribute patient load on resources. Lastly, mixed scheduling strikes a balance between block and spread-out scheduling by being more dispersed than block and more condensed than spread-out scheduling. Delays and early arrivals were accounted for via a delay function that adjusts patient arrival times by adding (or subtracting) some random amount of time to (from) appointment times.

**Table 1.**
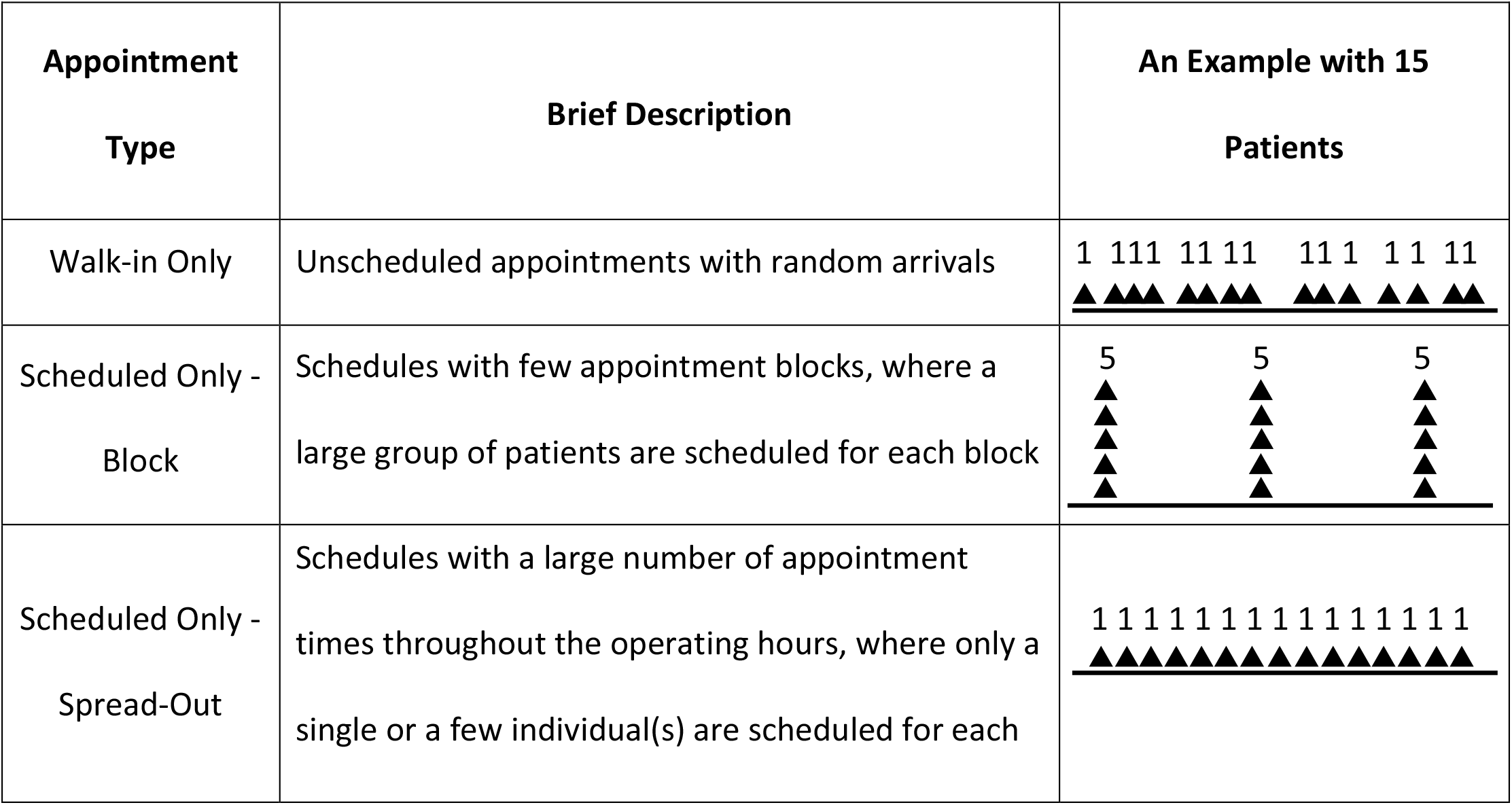
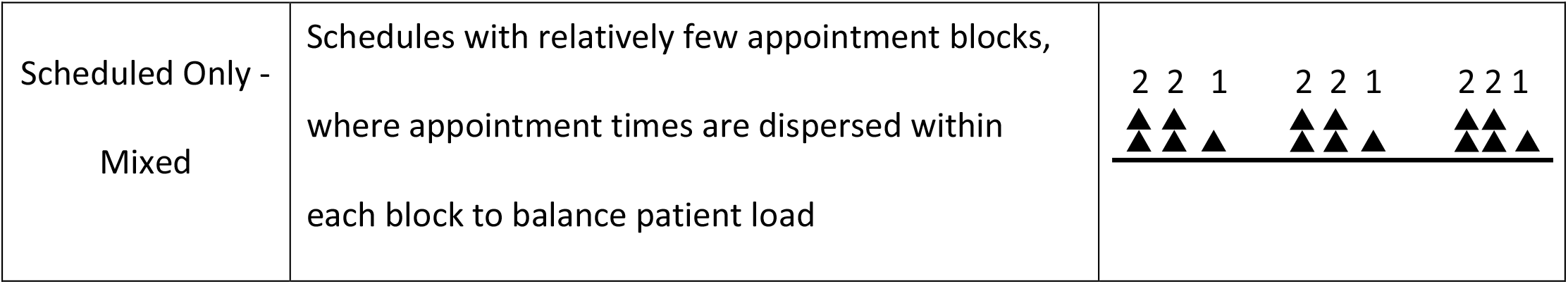
Different Appointment Types Considered in Our Analysis

| Appointment Type | Brief Description | An Example with 15 Patients |
| --- | --- | --- |
| Walk-in Only | Unscheduled appointments with random arrivals | 1 111 11 11 11 1 1 1 1 1<br>▲▲▲▲▲▲▲▲▲▲▲▲▲▲▲ |
| Scheduled Only - Block | Schedules with few appointment blocks, where a large group of patients are scheduled for each block | 5 5 5<br>▲▲▲▲▲▲▲▲▲▲▲▲▲▲▲ |
| Scheduled Only - Spread-Out | Schedules with a large number of appointment times throughout the operating hours, where only a single or a few individual(s) are scheduled for each | 1 1 1 1 1 1 1 1 1 1 1 1 1 1 1<br>▲▲▲▲▲▲▲▲▲▲▲▲▲▲▲ |

**Table 1.** Different Appointment Types Considered in Our Analysis
|  |  |  |
| --- | --- | --- |
| Scheduled Only -<br>Mixed | Schedules with relatively few appointment blocks,<br>where appointment times are dispersed within<br>each block to balance patient load | 2 2 1 2 2 1 2 2 1 |

### Monoclonal Antibody Treatment Process

The mAb treatment process in the decision-support tool starts with pre-infusion check-in (**Figure 1**). There are no capacity restrictions in the check-in area, as arriving patients are directed to wait outside when this area is fully occupied. Each check-in area staff serves a single patient at a time and performs clerical and clinical duties such as obtaining patient information and signatures for consent forms, verifying patient identification and health insurance, providing information about the treatment, and checking vitals. At the end of this process, clinical staff in walk-in facilities inform the pharmacy to initiate medication preparation. This step is bypassed for scheduled appointments, as medications are assumed to be prepared in advance for scheduled visits.

**Figure 1.**
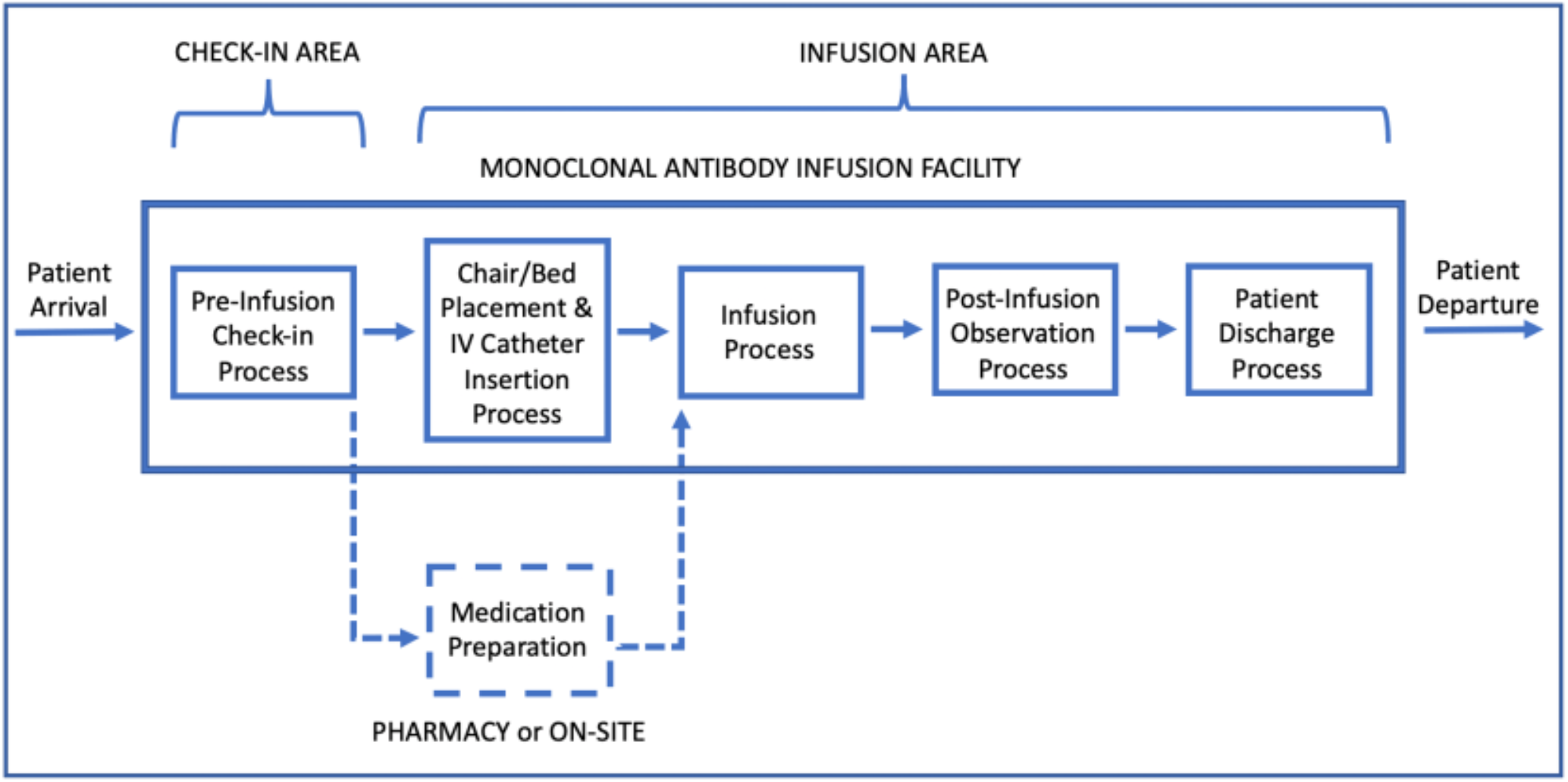
Patient Flow Diagram in a Monoclonal Antibody Infusion Facility

Following the waiting time after check-in, patients enter the infusion area and are directed to a treatment chair/bed. Consisting of nurses and allied health professionals (e.g., paramedics), the medical team in the infusion area prepare patients for infusion by conducting a medical examination, and inserting a peripheral intravenous (IV) catheter for drug administration. “Chair/Bed placement and IV catheter insertion” is followed by the infusion process. Infusion therapy requires a fixed amount of time, depending on medication, and is delayed when there is no medication readily available or if the medical care team is busy attending other patients.

The infusion process is followed by an observation period, during which patients remain seated and are monitored. Post-infusion observation period takes an hour but might be prolonged by concerns about a patient’s health status. Following the observation, the IV is removed and each patient goes through a discharge process, waiting for the final paperwork and the discharge consent from the physician in charge. Subsequently, patients depart the mAb infusion facility.

### Data Sources, Data Collection, and Simulation Parameters

The parameters and probability distributions used in simulation experiments reflect the data collected during observation of three U.S. medical centers implementing mAb infusions (**Table 2**). These parameters correspond to service durations and do not include preceding or succeeding waiting times. Data and descriptions for the generation of scheduled and walk-in arrivals and delays during the observation process are provided in the **Supplemental Material - Appendix A**.

**Table 2.**
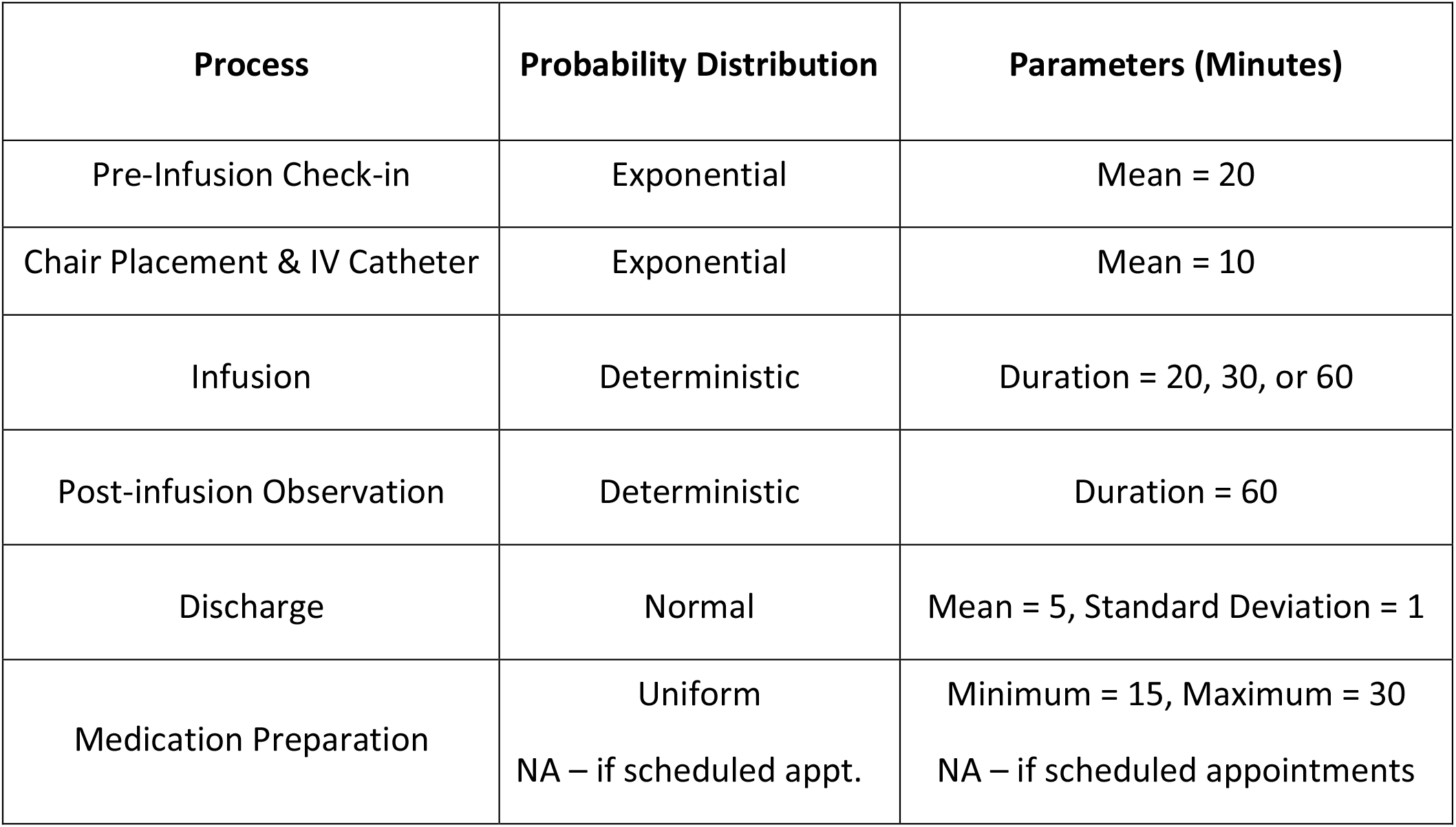
Probability Distributions and Parameters for mAb Treatment Sub-Processes

| Process | Probability Distribution | Parameters (Minutes) |
| --- | --- | --- |
| Pre-Infusion Check-in | Exponential | Mean = 20 |
| Chair Placement & IV Catheter | Exponential | Mean = 10 |
| Infusion | Deterministic | Duration = 20, 30, or 60 |
| Post-infusion Observation | Deterministic | Duration = 60 |
| Discharge | Normal | Mean = 5, Standard Deviation = 1 |
| Medication Preparation | Uniform<br>NA – if scheduled appt. | Minimum = 15, Maximum = 30<br>NA – if scheduled appointments |

### Discrete-Event Simulation (DES) Model

A simulation model was developed to analyze the COVID-19 mAb infusion therapy process and assess the performance of mAb infusion sites under various scenarios. Model inputs were as follows: (i) treatment bed/chair capacity, (ii) type of appointments, (iii) service hours, (iv) daily patient demand, (v) infusion duration, and (vi) staffing levels at check-in and infusion areas (**Table 3**). Model outputs were (i) average patient length of stay (LoS) with 95% confidence intervals, (ii) number and percentage of patients treated during service hours, and (iii) percentages of patients treated within three and four hours.

**Table 3.** Model Inputs for Discrete Event Simulation Model

| **Model Variables** | **Alternative Input Values** | **Number of Alternatives** |
| --- | --- | --- |
| Physical Capacity – Number of Treatment Beds/Chairs | 3 , 5 , 6 , 9 , 10 , 12 , 15 ,<br>20 , 30 | 9 |
| Appointment Type | Walk-in<br>Scheduled – Block<br>Scheduled – Mixed<br>Scheduled – Spread-Out | 4 |
| Operating Hours | 6 , 8 , 10 , 12 , 24 hour | 5 |
| Daily Patient Demand | 10 , 15 , 20 , 25 , 30 | 5 |
| Infusion Duration | 20, 30, 60 minutes | 3 |
| Check-in Area Staffing Levels | 1 , 2 , 3 | 3 |
| Infusion Area Staffing Levels<br>(Nurse) | 1 , 2 , 3 , 4 | 4 |
| Infusion Area Staffing Levels<br>(Allied Health Professional) | 0 , 1 , 2 , 3 , 4 | 5 |

Inputs for staffing levels were concentrated on the number of nurses and allied health professionals. Other roles, such as physicians or pharmacists, are also critical for infusion facilities, but were not observed to directly limit the capacity and patient flow. Based on on-site observations, a nurse was assumed to deliver concurrent care for up to five patients, whereas an allied health professional was assumed to simultaneously care for up to three patients. Model assumptions required at least one nurse to be present in infusion area. The type of clinical staff was assumed not to impact check-in tasks, where a single employee serves one patient at a time.

The simulated events for mAb infusion process were (i) patient arrivals, (ii) check-in service completion, (iii) service completion for chair/bed placement and IV catheter insertion, (iv) completion of infusion therapy, (v) end of post-infusion observation, (vi) patient departure following discharge process, and (vii) closure of the infusion clinic. Walk-in clinics required an additional event of medication preparation since they do not start preparing infusion medication before patient arrival and check-in. The model structure and status updates performed for these events are described in the **Supplemental Material - Appendix B**.

A verification analysis was conducted to confirm that the simulation model was consistent with the mAb therapy process and experiments were correctly implemented. Examining 162,000 simulated scenarios, we observed that patient LoS and other output metrics either improved or remained constant but never worsened when staffing or capacity levels were increased while other inputs were kept constant. Similarly, all output metrics improved, though not at the same magnitude, when infusion time was reduced from 60 minutes to 30 minutes and from 30 minutes to 20 minutes. Finally, all output measures in 40,500 walk-in scenarios worsened when medication preparation time range was increased from 15-30 minutes to 30-60 minutes. These verification analyses confirmed logical outcomes and that the simulation experiments were performed correctly.

Since mAb infusion for COVID-19 is a novel therapy, there are insufficient data to perform a comprehensive validation analysis. However, the accuracy of the generated estimates was assessed using expert face validation. Several feedback and pilot sessions were conducted with partner mAb sites and experts were asked to validate estimates for performance metrics. Using the web-based decision-support tool, they evaluated multiple different scenarios, and affirmed that estimated outputs were in line with their observations and expectations.

### Web-Based Decision-Support Tool: mAb Infusion Process Calculator

Utilizing simulation results, a web-based calculator (**Figure 2)** was created to assist capacity planning and resource allocation decisions for mAb infusion sites. At the website, users provide inputs for the main variables of a mAb infusion process (**Table 3**) and the outputs of the corresponding scenario are displayed to the user. There are 162,000 scenarios that users can investigate. All simulation experiments were performed in advance and saved on the website to provide instant feedback to users.

**Figure 2.**
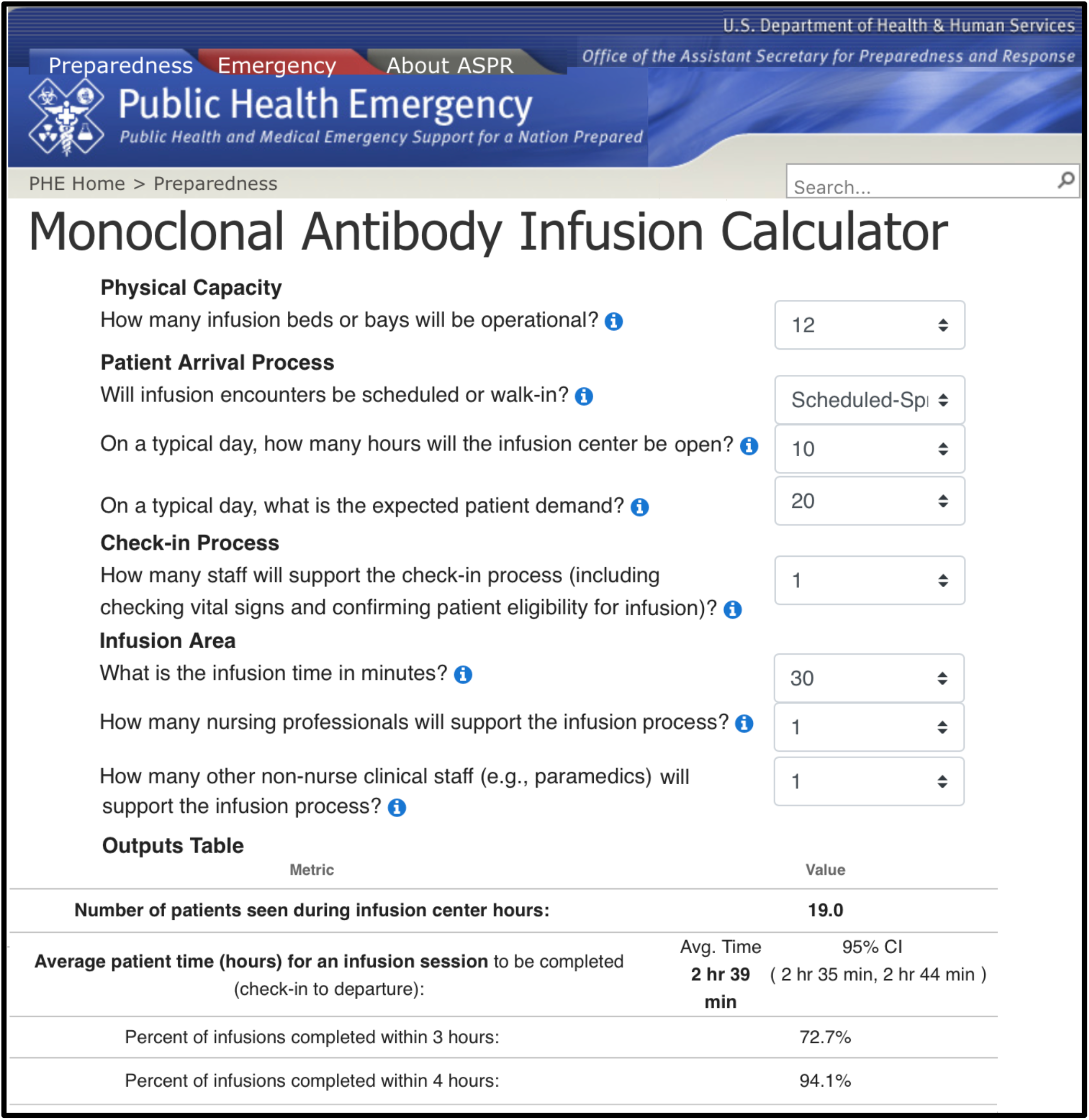
User Interface of the Web-Based mAb Infusion Process Calculator

In addition to simulation outputs, the web-based calculator also displays two plots for each selected scenario (**Figure 3**). The first plot depicts the impact of increasing or decreasing the number of infusion nurses on the total number of patients that can be served throughout a day.

**Figure 3.**
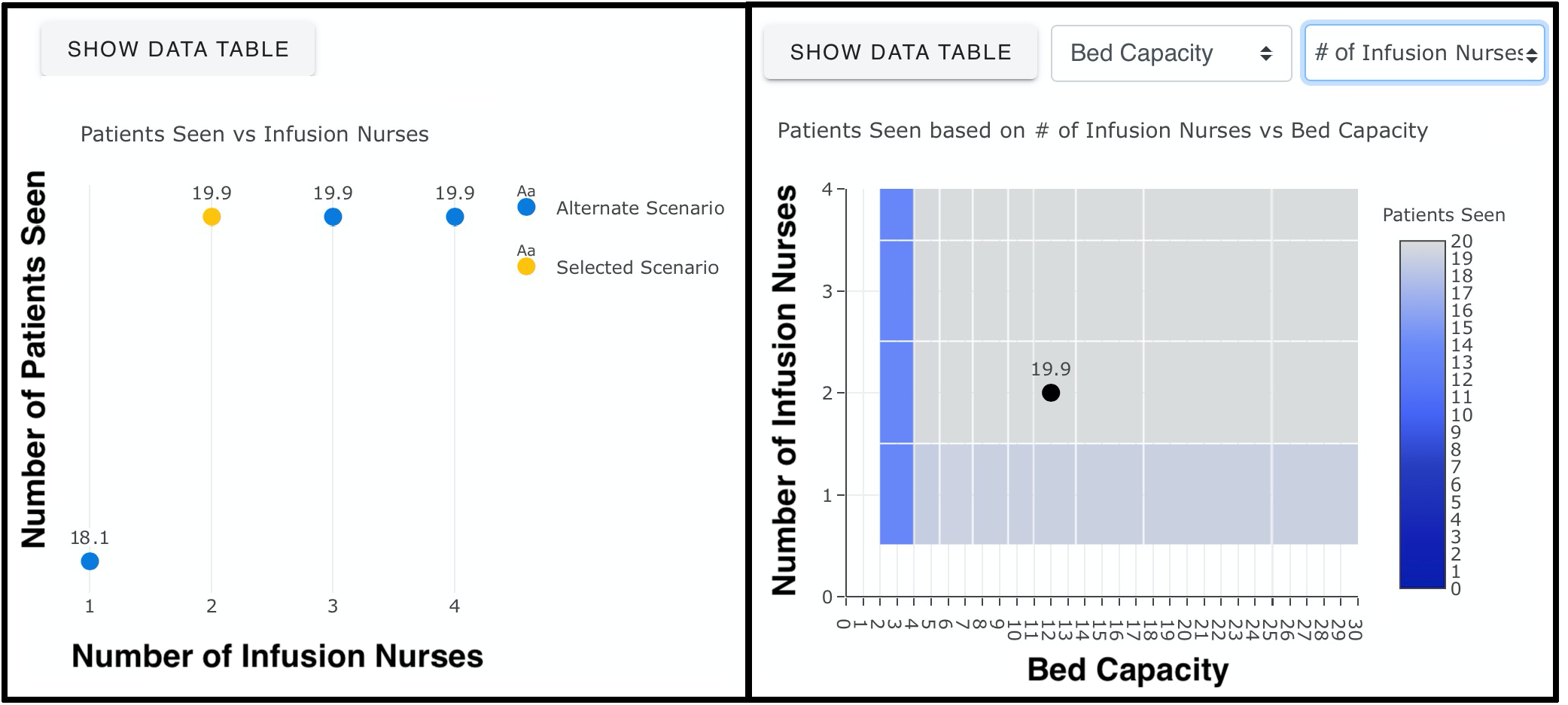
Graphs Generated by the Web-Based mAb Infusion Process Calculator

The second plot shows the effect of any two variables chosen by the user (e.g., number of infusion nurses and bed capacity) on the number of patients treated. Together, the graphs help users assess the value of changing staffing levels for a given scenario, examine how added resources lead to improvements or diminishing returns, and identify the bottlenecks of the simulated scenario. Further, these graphs could be used to determine the minimum number of resources required to meet desired service targets in terms of daily patient throughput.

## 3. Results

A total number of 162,000 different scenarios were evaluated by simulation experiments. Each infusion duration considered in the analysis (namely, 20-, 30-, and 60-minute) corresponded to 54,000 different cases, where walk-in scenarios and different scheduling protocols each made up one fourth (i.e., 13,500) of these cases (**Supplemental Material - Appendix C**). All results can be accessed via the web-based calculator hosted at *https://www.phe.gov/emergency/mAbs-calculator/Pages/default.aspx*.

Based on 162,000 scenarios considered, various analyses can be performed to generate insights about different aspects of the mAb infusion process. To demonstrate the utility of the web-based calculator and shed light onto important planning and research questions for mAb infusion sites, we focused on four main areas: (i) the impact of scheduling on performance metrics, (ii) the effect of infusion duration on patient LoS, (iii) the role of medication preparation duration for walk-in encounters, and (iv) when and where to add additional staff to improve overall performance.

### Impact of Scheduling on Performance Metrics

To assess the impact of scheduling, we compared 13,500 walk-in scenarios with 60-minute infusion times to their 13,500 spread-out scheduling counterparts. Excluding 3,457 scenarios having < 10 minutes gap in LoS difference, we analyzed 10,043 scenarios with a non-negligible difference (i.e., ≥ 10 minutes gap) between LoS durations when spread-out scheduling was compared to walk-ins. Among these 10,043 scenarios, where the LoS difference were non-negligible, scheduled appointments had shorter LoS averages in 7,286 (72.5%) scenarios, and walk-in clinics achieved lower patient LoS in 2,757 (27.5%) cases.

Our analysis revealed that *traffic intensity*†, a measure of the average occupancy of a service area, could play a key role in identifying when scheduling is the most and least beneficial. We observed that when the traffic intensity level was low to medium in infusion and check-in areas, indicating that the patient load on the mAb infusion site was manageable, scheduling yielded better outcomes and was an effective means for providing timely mAb treatments. In particular, there were 7,916 scenarios when the traffic intensity of the check-in process was < 2.94 (i.e., medium), and scheduling resulted in lower LoS in 7,247 (91.5%) of these cases. Similarly, LoS averages were shorter with scheduling in 7,068 (90.2%) out of 7,839 scenarios when the traffic intensity in the infusion area was below 0.35 (i.e., low). However, scheduling had a limited impact on reducing LoS when the traffic intensity was high in either service area (**Table 4**). Compared to walk-in clinics, implementation of scheduled appointments achieved no improvement on LoS durations when traffic intensity was ≥ 3.86 in the check-in and ≥ 0.43 in the infusion area.

**Table 4.**
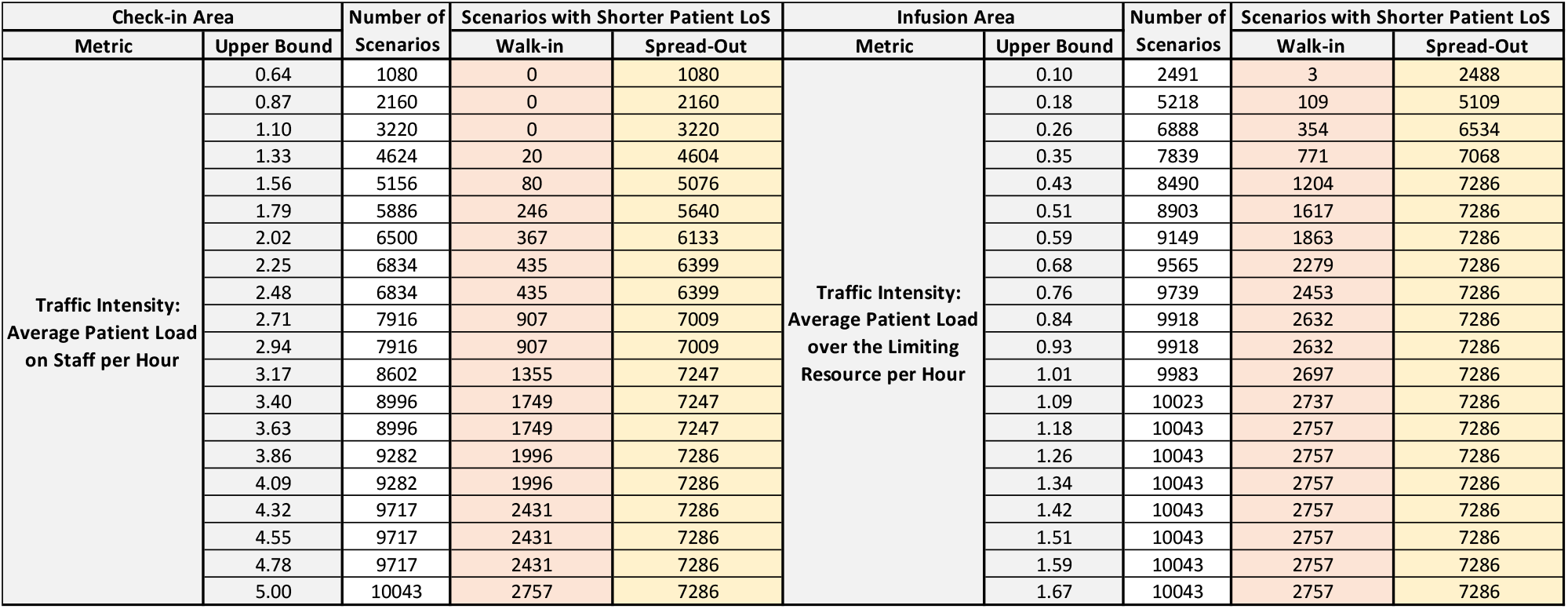
Number of Scenarios with Shorter LoS under Walk-ins and Spread-Out Schedules

### Effect of Infusion Time on Patient Length of Stay

Infusion times ranged from approximately 20-60 minutes. To investigate the effect of infusion time on patient LoS, we compared the average LoS outputs of 54,000 scenarios that had 30-minute infusion time to their 108,000 counterparts with 20- and 60-minute-long infusions. The distributions of LoS difference when 30-minute infusions compared to other two alternatives are presented in **Figure 4**.

**Figure 4.**
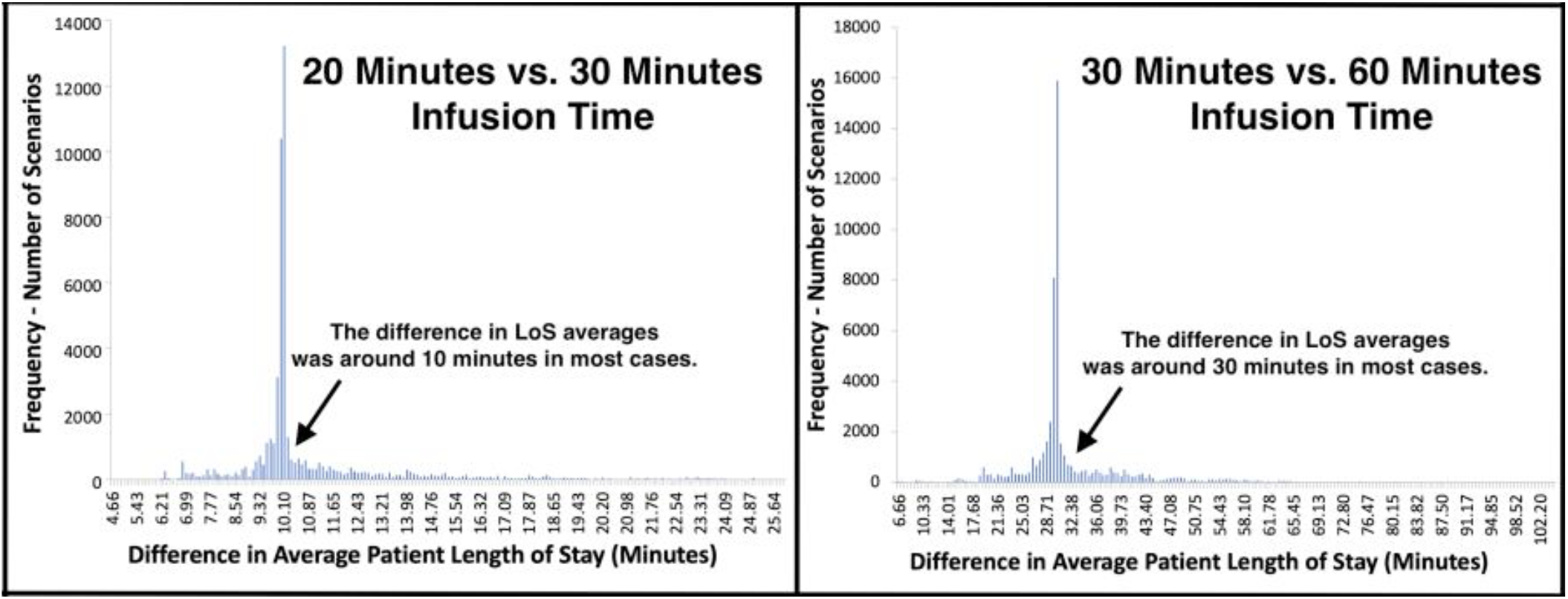
Distribution of LoS Difference when Different Infusion Times were Compared

The differences in LoS averages centered around the amount of change in infusion times, which was a 10-minute decrease and a 30-minute increase for 20- and 60-minute infusions, respectively (**Figure 4**). Yet, noticeably higher/lower changes in patient LoS were observed for a fair number of comparisons. In particular, 4,000 (7.4%) scenarios achieved > 45 minutes reduction in LoS when infusions took 30 minutes instead of 60 minutes. Similarly, 4,530 (8.4%) scenarios had > 15 minutes difference in LoS when 20-minutes infusions were compared to 30 minutes infusions.

For the comparisons having significantly higher/lower change in LoS, queueing theory concepts “traffic intensity”, measuring the average occupancy of a service system, and “stability”, indicating whether its traffic intensity is at manageable levels or not, were helpful to explain this phenomenon [12, 13]. The improvements observed in LoS were noticeably higher when reducing infusion duration led an unstable infusion site to experience manageable traffic intensity and become stable. This was because, by achieving stability, the whole service system functioned more efficiently, waiting times between consecutive services were reduced, and as a result, total reductions in LoS were higher than the change in infusion time. Similarly, the performance of a stable infusion site significantly worsened following an increase in infusion time when this change caused its traffic intensity to rise above manageable levels.

### Role of Medication Preparation Duration for Walk-In mAb Clinics

At walk-in clinics, the preparation of mAb solution was performed after patient check-in rather than beforehand. In general, this might cause delays in infusion therapy. To quantify the effect of medication preparation on patient LoS, we conducted additional experiments, where medications were prepared within 30-60 minutes, and compared them to the baseline scenarios of 15- to 30-minute-long medication preparation time. The difference between LoS averages mostly accumulated near 22.5 minutes, which was the average difference between 15-30 (mean time 22.5) and 30-60 (mean time 45.0) minute preparation times (**Figure 5**). Changing the duration made less impact on LoS when physical capacity was low (i.e., number of beds ≤ 6) or patient load was high (i.e., ≥ 15 patients in ≤ 12 service hours). For those infusion sites, increasing bed capacity or staffing could be more beneficial than preparing infusion medications faster.

**Figure 5.**
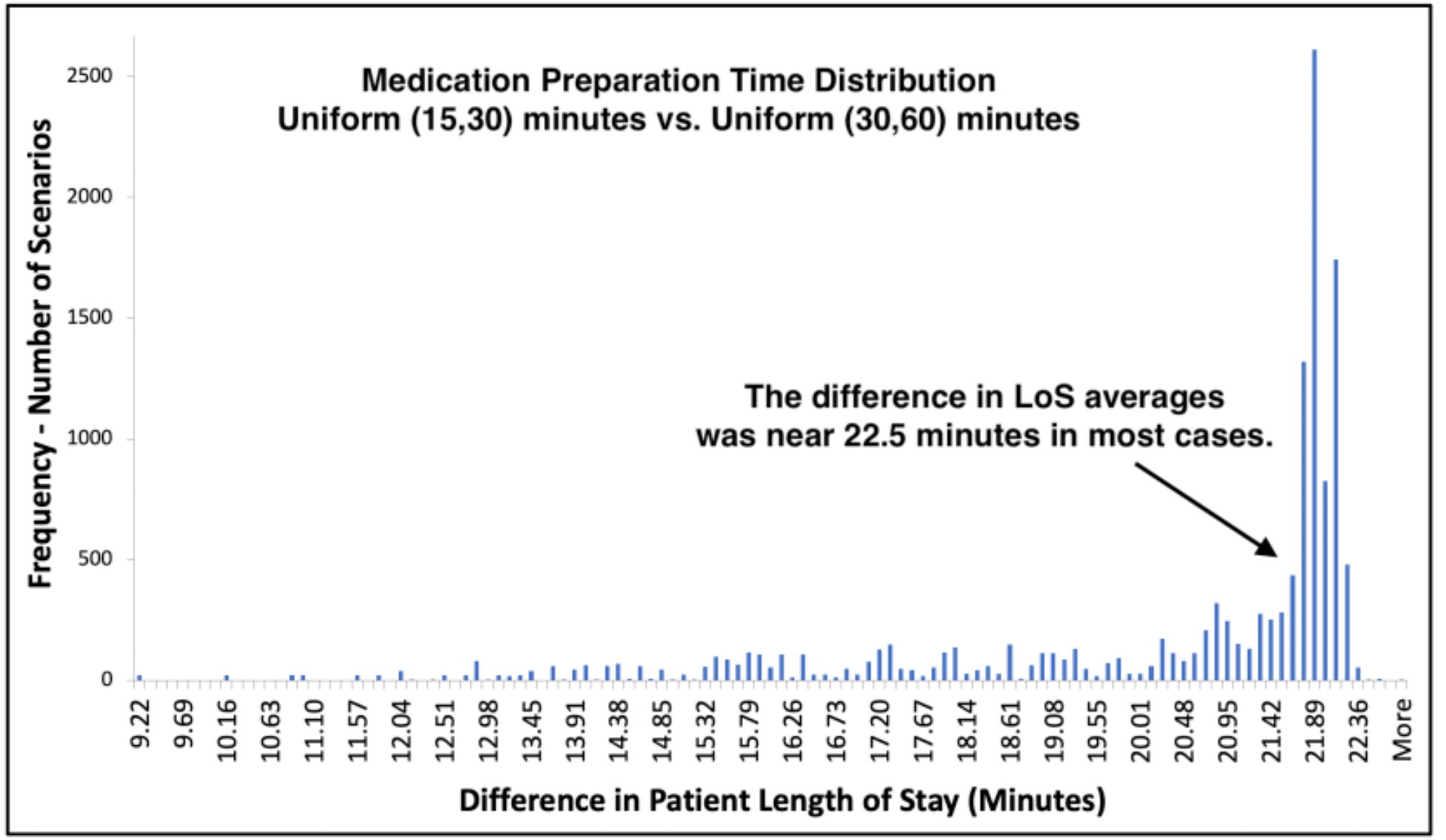
Distribution of LoS Difference under Different Medication Preparation Times – Comparison of 13,500 Walk-in Scenarios with 60 Minute Infusion Time

### Staff Adjustments to Improve Overall Performance of a mAb Infusion Site

Simulated scenarios with 60-minute infusion times (n = 54,000) were examined separately for each scheduling type (n= 13,500) to assess the impact of adding staff members under different roles (**Figure 6**). To simplify the analysis, the scenarios, where infusion teams consisted of only nurses (n = 2,700), were considered. The cases for which the limiting factor was physical capacity rather than staffing were excluded. Consequently, a total of 650 scenarios were identified, for which staffing levels in check-in and infusion areas could both be increased. For these 650 scenarios, a staff was exclusively added to only one area and then, LoS averages were compared.

**Figure 6.**
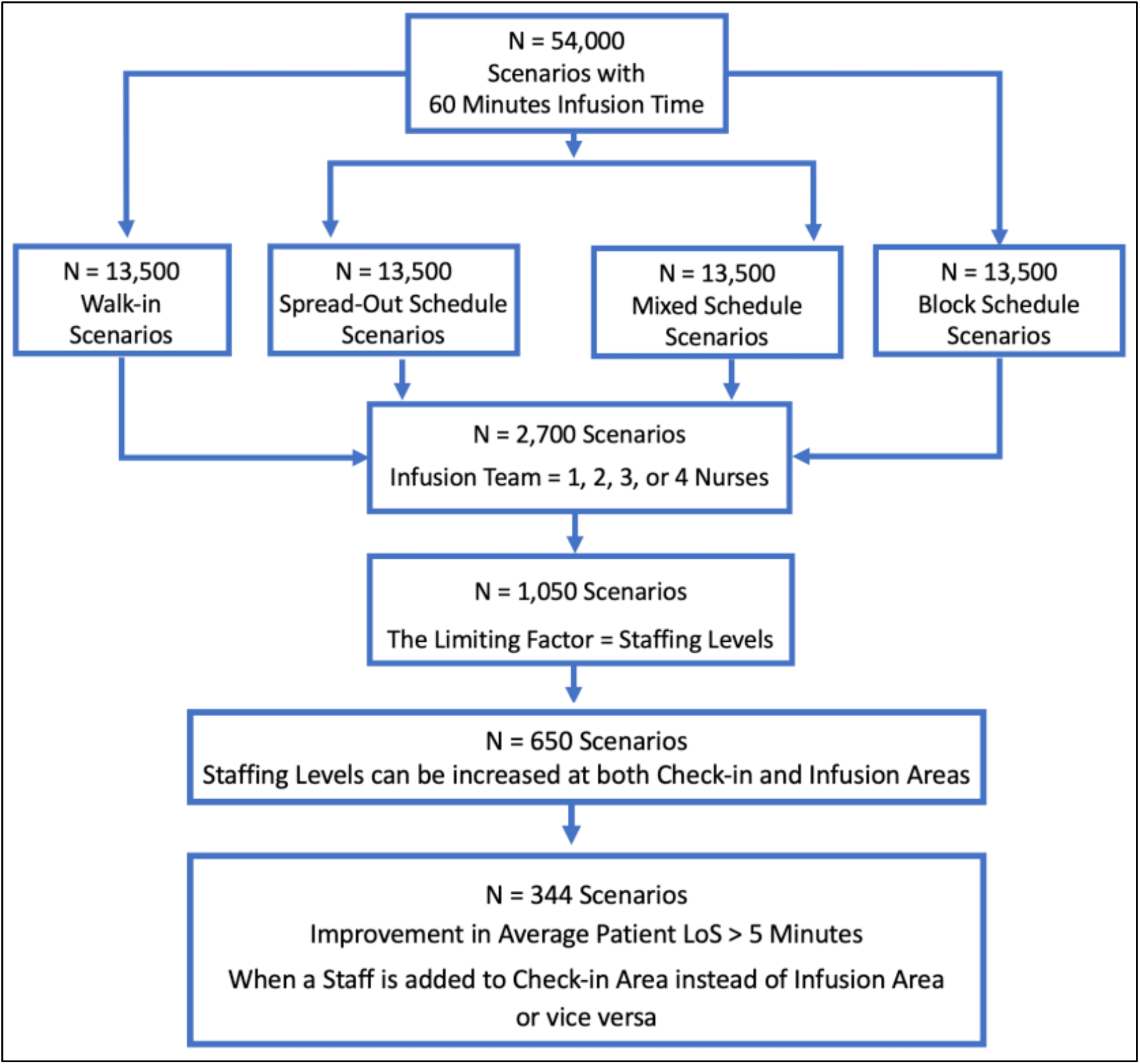
CONSORT Diagram - The Number of Scenarios Included in Staff Adjustment Analysis

In 344 scenarios, the reduction in LoS achieved by adding a staff to an area were > 5 minutes shorter than the other alternative (**Figure 6**). In 212 (62%) scenarios, adding an infusion nurse resulted in a larger decrease to LoS (mean 34 minutes). In the remaining 132 (38%) scenarios, where adding a check-in staff member was more beneficial (mean 24 minutes), physical capacity was ≥ 12 beds, the number of infusion area nurses was ≥ 2, and check-in staff were always originally fewer. These similarities in 132 scenarios suggest that, for medium-to-large size infusion sites having more than ten or eleven beds, providing support to check-in area might be more beneficial when there is a single check-in staff member and more than one infusion nurse.

## 4. Discussion

Based on data collected from three U.S. COVID-19 mAb infusion centers, we conducted simulations allocating personnel and other resources in mAb infusion sites and translated the results into a web-based decision-support tool. The simulation experiment results and our analyses led to several important findings. In particular, despite nursing staff shortages often being perceived as the main barrier, this study identified that other factors were equally important for the overall performance of a mAb site. Frequently, patient LoS was extended due to other factors such as physical capacity, staffing levels at check-in, or the duration of infusion process, even with low nursing levels. These findings suggest that decisions should be made carefully for all key components of a mAb infusion process to provide timely service and improve staffing and capacity efficiency.

Regarding the value of appointment scheduling, it is common to assume scheduling would yield better performance as it allows planning and preparation for upcoming patients beforehand. Yet, this analysis revealed that scheduling appointments was not necessarily beneficial when daily patient volumes were above a level that a mAb site can effectively manage. In those situations, patient load on resources should be reduced by either extending business hours or increasing the levels of the limiting resource.

Achieving a reduction in the duration of a critical service, such as infusion or medication preparation time, did not always lead to significant improvements. In particular, when an infusion site continued to experience high traffic intensity resulting from high patient volumes, short business hours, or low capacity and staffing levels, improvements in patient LoS were minimal. However, when traffic intensity was only slightly above manageable levels, shortening the duration of a medical service (e.g., infusion time) achieved stability and made a significant impact for the operational performance of mAb sites. These results suggest that the infusion sites experiencing patient traffic that is only marginally higher than their physical and staff capacity could benefit the most from shortened service durations, which, for instance, can be achieved by using subcutaneous route for mAb administration instead of IV route.

The demand and need for mAb infusions will vary across communities and over time. Overall, the results from this study suggest that health systems in the U.S. can effectively provide mAb treatments under different settings even with relatively low investments for physical resources and staff. This is not to say that the launch of a mAb site is without barriers, but that this service can be made available to a community and reduce hospitalizations and COVID-19 deaths without a heavy burden on resources. The key, here, is to correctly identify the bottlenecks of the mAb infusion process in the corresponding setting and make the right adjustments to efficiently use limited resources. By analyzing mAb infusions under different conditions and resource allocations, this study can offer guidance for the optimal planning and implementation of mAb infusion sites, which can have a considerable impact in regional COVID-19 response efforts.

This study was not free of limitations. First, the process model aligned with field observations for several sites and was validated by focus groups with those same sites. Hospitals may choose to implement mAb infusions in a different setting. Second, simulations relied on several assumptions for service times, nurse-to-patient ratio in infusion area, and arrival patterns under different scheduling protocols. Differences in these assumptions could impact simulation results. Third, despite considering the majority of the practical scenarios, the web-based decision-support tool is not exhaustive of all possible cases. Accordingly, users will not always be able to select the exact model input values corresponding to the scenario(s) they desire to investigate. Finally, in addition to IV infusions, mAb treatments have also now been approved as subcutaneous injections, which were not the focus of our study.

## 5. Conclusion

This effort used a simulation approach to create a capacity and staffing planning tool to support adoption and spread of mAb therapy for COVID-19. The objective was to better understand staffing, physical resources, and other requirements for providing timely mAb infusion services with efficient resource use. Considering different staffing and capacity levels, scheduling protocols, patient demand, service hours, and infusion durations, 162,000 scenarios were evaluated via simulation experiments. A web-based decision-support tool was created to allow decision-makers, researchers, and other users to easily access the results, investigate operational performance of mAb infusion sites under different conditions, and generate managerial insights for existing and future infusion sites. Given that mAb treatments are expected to continue being an important instrument for the management of COVID-19, the web-based calculator introduced in this study could significantly contribute to pandemic control, planning, and preparation efforts.

## Data Availability

Real data were collected from three monoclonal antibody infusion sites.
The collected data were about the service times of each service process (e.g., how long an infusion takes) and did not include any information about individual patients.

## 6. Acknowledgements

This study was supported by the U.S. Department of Health and Human Services’ Office of the Assistant Secretary for Preparedness and Response. The findings and conclusions in this paper are those of the authors and do not represent the official position of any government or private organization.

## Supplemental Material

### Appendix A – Data and Description for Generating Patient Arrival Times and Adverse Events

For walk-in appointments, we took a two-step approach to generate patient arrivals. First, utilizing an emergency department data from a large-scale hospital, we derived an empirical probability density function for the number of arrivals, measuring the likelihood of a patient arrival for each one-hour interval during a day. Next, for a given total number of arrivals pre-specified by the scenario to be simulated, we used this empirical probability density function to determine how many of these patients to arrive at each one-hour slot during the service hours. Once the number of arriving patients for each one-hour interval was determined, we then employed a uniform probability distribution for every one-hour interval to generate the arrival times of the walk-in patients. For scheduled appointments, appointment times were determined by the scheduling type. Yet, we also adjusted the patient arrival times around pre-scheduled appointment times by using normally distributed a random variable to account for delays and early arrivals. The probability distributions and parameters for walk-in arrivals and late and early arrival adjustments for scheduled arrivals are provided in **Table A1**.

**Table A1.** Parameters for Early or Late Arrivals and Probability Function for Walk-in Arrivals

| Process | Probability Distribution | Parameters (Minutes) |
| --- | --- | --- |
| Early or Late Arrival to Scheduled Appointments | Normal | Mean = 7.5,<br>Standard Deviation = 2 |
| Walk-in Arrival | Empirical | Prob. = 0.0517 for 9.00 - 9.59 am<br>Prob. = 0.0623 for 10.00 - 10.59 am<br>Prob. = 0.0617 for 11.00 - 11.59 am<br>Prob. = 0.0613 for 12.00 - 12.59 pm<br>Prob. = 0.0576 for 1.00 - 1.59 pm<br>Prob. = 0.0564 for 2.00 - 2.59 pm<br>Prob. = 0.0565 for 3.00 - 3.59 pm<br>Prob. = 0.0561 for 4.00 - 4.59 pm<br>Prob. = 0.0560 for 5.00 - 5.59 pm<br>Prob. = 0.0560 for 6.00 - 6.59 pm<br>Prob. = 0.0574 for 7.00 - 7.59 pm<br>Prob. = 0.0514 for 8.00 - 8.59 pm<br>Prob. = 0.0464 for 9.00 - 9.59 pm<br>Prob. = 0.0397 for 10.00 - 10.59 pm<br>Prob. = 0.0287 for 11.00 - 11.59 pm<br>Prob. = 0.0277 for 12.00 - 12.59 am<br>Prob. = 0.0217 for 1.00 - 1.59 am<br>Prob. = 0.0185 for 2.00 - 2.59 am<br>Prob. = 0.0175 for 3.00 - 3.59 am<br>Prob. = 0.0160 for 4.00 - 4.59 am<br>Prob. = 0.0159 for 5.00 - 5.59 am<br>Prob. = 0.0187 for 6.00 - 6.59 am<br>Prob. = 0.0248 for 7.00 - 7.59 am<br>Prob. = 0.0400 for 8.00 - 8.59 am |

Both for walk-in and for scheduled appointments, occasionally, we observed prolonged post-infusion observation times. Despite being rare, the delay resulted from several reasons including concerns about the observed patient’s health status, medical equipment failure, or minor adverse events such as bleeding after IV catheter removal. The frequency of a prolonged observation was 5% and the duration of an increase in post-infusion observation service was 15, 30, 45, or 60 minutes with probabilities 0.50, 0.25, 0.20, and 0.05, respectively (See **Table A2**).

**Table A2.** Parameters for Prolonged Post-Infusion Observation Process

| Process | Probability Distribution | Parameters (Minutes) |
| --- | --- | --- |
| Prolonged Observation | Discrete | Probability of Occurrence = 0.05<br>Duration of Delay = 15, 30, 45, or 60<br>Duration Prob. = 0.50, 0.25, 0.20, 0.05 |

### Appendix B - The Simulation Events and Main Steps of The Simulation Algorithm

From patient arrival to departure, the overall mAb treatment process typically consists of pre-infusion check-in, chair placement and IV catheter insertion, infusion, post-infusion observation, and discharge steps. For walk-in appointments, these processes are accompanied by medication preparation process, performed either on-site or at an external pharmacy, whereas medication preparation is assumed to be completed in advance for scheduled appointments. Among these activities, the discrete-event simulation algorithm identifies the next event at each iteration, performs system status updates corresponding the next simulation event, advances the simulation clock, and then performs the next iteration by repeating this simulation logic until the closure time of the mAb facility. The events that are modeled and simulated by the discrete-event simulations are briefly described as follows:

#### Patient Arrival

In the check-in area, the patients are served based on first-come-first-served rule. Accordingly, a newly arrived patient begins her check-in process if there is an idle staff member in the check-in area. Otherwise, she joins the queue and waits for her service either in the check-in area or outside (e.g., in her car).

#### Check-in Process Completion

Following the completion of the pre-infusion check-in service, each patient is admitted to the infusion area given the physical capacity permits and the infusion area clinical staff, consisting of nurses and allied health professionals (e.g., paramedics), are available to initiate her bed/chair placement and IV catheter insertion process. Yet, when all of the treatment beds/chairs are occupied or the clinical personnel in the infusion area are busy with serving other patients, patients continue to stay in the check-in area and wait. Meanwhile, the check-in process of another patient can be initiated.

#### Chair/Bed Placement and IV Catheter Insertion Completion

Once a patient is placed on a treatment bed/chair and her IV catheter is inserted, the patient becomes ready for her mAb infusion therapy given her medication is also available. At this point, the medical team decides whether to initiate the infusion process of this patient or bed/chair placement and IV catheter insertion of another patient depending on the longest waiting line. Note that infusion process cannot begin if there is no medication available and similarly, bed/chair placement process cannot be initiated if there is no idle treatment bed/chair and available medical personnel. When neither process can be initiated, a member of the infusion team performs discharge tasks for one of the patients waiting for their discharge process to be completed before their departure.

#### Infusion Process Completion

The infusion process is automated, takes a fixed amount of time (e.g., 20, 30, or 60 minutes) depending on the medication, and is performed under the supervision of the infusion area medical team. Once it is completed, the one-hour long observation process begins for the patient. At the same time, the infusion team begins to discharge a patient waiting for her departure or initiates bed/chair placement or infusion process of another patient depending on the queue length and availability of a treatment bed/chair or infusion medication.

#### Observation Completion

Following the post-infusion observation process, a patient joins the discharge process queue and begins waiting for his discharge procedure to be initiated. Meanwhile, the infusion team initiates the discharge process for the head of the waiting line patient and continue to supervise other patients in the infusion area.

#### Discharge Completion

Discharge is the last process of the overall mAb infusion therapy, following which patients depart the mAb infusion center. Accordingly, after each discharge completion, a treatment bed/chair becomes available for the use of another patient. The medical team in the infusion area either admits a new patient to the infusion area by starting that patient’s bed/chair placement process or initiates the infusion process for a different patient. This choice is made based on the largest waiting queue length between bed/chair placement and infusion processes, where the lack of medication to be administered delays the infusion process.

#### Medication Preparation Completion

For scheduled appointments, the medication preparation is assumed to be performed in advance. For walk-in appointments, the completion of the check-in process initiates medication preparation either on-site or at an external pharmacy. Once a medication is prepared, it is sent to the infusion area and becomes available to be administered. If there is a patient waiting for her infusion process due to the lack of a medication, the newly prepared medication enables the initiation of the infusion process for this patient.

### Appendix C - The Structure of The Discrete-Event Simulation Model

The programming code for the discrete-event simulation model consists of two major parts: the main code and simulation function. By retrieving model inputs an excel document, the main code specifies the values of the model variables for the scenario of interest to be simulated such as the number of treatment beds, appointment type, and facility service hours. It also defines the probability distributions and parameters for the stochastic and deterministic processes (e.g., exponential distribution with mean 20 minutes for check-in process) and uses them to execute random number generations for every process from patient arrival to departure. Then, the main code forwards these inputs to the simulation function, commands it to run the scenario of interest with a pre-determined number of replications, records the summary statistics with performance measures, and reports the model outputs to the user for the evaluated scenario.

Discrete-event simulation experiments are performed by the second component of the programming code: the simulation function. A fixed number of replications are independently run by the simulation function for all of the scenarios to be simulated. For each replication of a scenario, the simulation code initiates its execution at the opening time of the mAb infusion site. Then, the code identifies the next event, updates the system status depending on the (changes the) next event (causes), collects statistics, and forwards the simulation clock to the time of the next event in this respective order. This process sequence is iterated by the simulation function until the closure time of the mAb infusion site, which marks the termination time of the current replication. Subsequently, the simulation function moves forward to running the next replication for the same scenario and reports the summary statistics together with performance outputs to the main code when all of the replications are completed for the given scenario.

Employing the discrete-event simulation model and utilizing the scheme described in this paper, 162,000 different scenarios were assessed by simulation experiments. The number of scenarios corresponding to different infusion durations and scheduling types are reported in **Table C**.

**Table C.** The Distribution of Simulated Scenarios

| Infusion Duration | Total Number of Scenarios | Scheduling Type | Number of Scenarios |
| --- | --- | --- | --- |
| 20 minutes | 54,000 | Walk-in | 13,500 |
|  |  | Spread-Out | 13,500 |
|  |  | Mixed | 13,500 |
|  |  | Block | 13,500 |
| 30 minutes | 54,000 | Walk-in | 13,500 |
|  |  | Spread-Out | 13,500 |
|  |  | Mixed | 13,500 |
|  |  | Block | 13,500 |
| 60 minutes | 54,000 | Walk-in | 13,500 |
|  |  | Spread-Out | 13,500 |
|  |  | Mixed | 13,500 |
|  |  | Block | 13,500 |

## Footnotes

† Traffic intensity estimates the average patient load on each treatment bed (staff member) per hour by multiplying service duration (e.g., check-in time) with the average number of arrivals per hour divided by the maximum capacity (service coverage) provided by treatment beds (staff).

